# ESUS-AI: a machine learning framework to estimate the most likely embolic source in embolic stroke of undetermined source

**DOI:** 10.64898/2026.02.04.26345615

**Authors:** A Bonura, J Juega, C Meza, J Kühne Escolà, M Muchada, M Rubiera, M Olivé, M Requena, M Rodrigo, N Rodriguez-Villatoro, D Rodriguez-Luna, F Rizzo, G Fiore, R Simonetti, N Brunelli, R Rodriguez, J Francisco, G Colangelo, M Ribó, C Molina, J Pagola

## Abstract

**Background and Purpose:** Embolic stroke of undetermined source (ESUS) emains a major diagnostic challenge in vascular neurology, as a substantial proportion of patients lack an identifiable embolic source despite standardized diagnostic workup. The failure of empiric anticoagulation strategies highlights the need for individualized, mechanism-oriented risk stratification. We aimed to develop a machine learning–based framework to estimate the most likely embolic source in ESUS using routinely available clinical data.

**Methods:** We retrospectively analyzed consecutive ESUS patients admitted to the Stroke Unit of Vall d’Hebron Hospital between 2020 and 2024. Three supervised machine learning models (XGBoost, Random Forest, and regularized logistic regression) were trained to independently predict the presence of left atrial enlargement (LAE), left ventricular dysfunction or akinesia (LVD), and complex aortic plaques (AP), based on demographic, clinical, laboratory, and imaging variables available at diagnosis. Model interpretability was assessed using permutation importance and SHAP analyses.

**Results:** Among 1,741 ESUS patients (mean age 71.5±14.6 years; 48.3% women), LAE was present in 40.5%, AP in 11.0%, and LVD in 6.5%. XGBoost achieved the best overall performance across targets (PR-AUC: 0.71 for LAE, 0.29 for AP, 0.44 for LVD).

Distinct and biologically coherent risk profiles emerged. LAE was driven by older age, elevated NT-proBNP, higher stroke severity, and a non-linear association with cholesterol. AP was associated with advanced age and traditional vascular risk factors. LVD showed a cardiomyopathic pattern characterized by elevated NT-proBNP, younger age, male sex, and severe strokes.

**Conclusions:** A machine learning–based approach can provide probabilistic, mechanism-oriented stratification in ESUS, capturing non-linear interactions among routinely available variables. This framework may support clinicians in prioritizing targeted diagnostic pathways and tailoring secondary prevention strategies, pending external validation.

## Introduction

Despite accounting for nearly one quarter of all ischemic strokes^1,2^, Embolic Stroke of Undetermined Source (ESUS) remains a major diagnostic and therapeutic challenge in vascular neurology. ESUS frequently lacks a clearly identifiable cause such as atrial fibrillation detected after stroke (AFDAS), severe atherosclerosis, or other well-known etiologies, limiting the ability to tailor secondary prevention. Randomized clinical trials have shown that empirical anticoagulation in ESUS does not reduce stroke recurrence compared with antiplatelet therapy, underscoring the need for a more individualized, mechanism-based approach to prevention^3–5^.

According to the latest ESUS consensus, the most probable underlying mechanisms include left atrial disease, left ventricular dysfunction, and complex aortic atheroma. Rupture of aortic plaques may generate emboli, whereas left atrial enlargement and left ventricular akinesia reflect atrial or ventricular cardiomyopathies that promote blood stasis and thrombus formation^6,7^ ^8–10^ ^11,12^. These mechanisms represent distinct pathophysiological entities, yet they are frequently associated with overlapping clinical characteristics and shared vascular risk factors, which contribute to stroke recurrence but hinder reliable etiological attribution^13^.

In clinical practice, commonly used diagnostic tools—such as transthoracic echocardiography, prolonged cardiac rhythm monitoring, or aortic computed tomography—can identify some embolic sources, but their overall diagnostic yield remains limited. More sensitive investigations, including transesophageal echocardiography or implantable loop recorders, are invasive, resource-intensive, and not systematically available, resulting in a substantial proportion of ESUS patients in whom the embolic mechanism remains unresolved. As a consequence, preventive strategies are often implemented without a clear mechanistic rationale.

This diagnostic uncertainty reflects the complexity of ESUS, in which multiple clinical, imaging, and biological markers contribute to embolic risk through non-linear and interdependent relationships. Traditional statistical models, which assume linear and independent associations, are limited in their ability to integrate such multidimensional information and to weigh competing diagnostic hypotheses when direct evidence is incomplete.

In this context, machine learning approaches offer a complementary framework by jointly modeling heterogeneous variables and capturing latent interaction patterns that may reflect distinct pathophysiological substrates rather than single dominant risk factors^14^ ^15^. Rather than replacing definitive diagnostic criteria, ML-based models can support etiological inference in ESUS by prioritizing the most plausible underlying mechanism and guiding more targeted diagnostic and preventive strategies.

The aim of this study was to use a machine learning–based approach to identify clinical variables associated with the main suspected ESUS mechanisms—left atrial enlargement, left ventricular akinesia/dysfunction, and complex aortic plaques—and to stratify patients into distinct mechanistic risk profiles. By providing a probabilistic etiological stratification at the time of diagnosis, this approach is intended to support clinicians in prioritizing diagnostic pathways and tailoring secondary prevention strategies, rather than replacing established diagnostic criteria.

## Methods

Clinical data were retrospectively obtained from a dataset of patients fulfilling the ESUS admitted to the Stroke Unit of Vall d’Hebron University Hospital (Barcelona, Spain) between January 2020 and August 2024 included in the Catalan Stroke Registry, a prospective database audited by the Government of Catalonia^16^. This study was conducted in accordance with STROBE and TRIPOD – AI.

### Inclusion and exclusion criteria

All the patients included met the following criteria: a) confirmed diagnosis of ESUS according to international guidelines: non lacunar infarction with absence of stroke etiology b) availability of complete demographic and clinical data including: laboratory tests, computed tomography (CT) assessment, and CT angiography (CTA); c) Cardiologic workup with transtoracic echocardiography (TTE) or transesophagic echocardiography (TEE) and at least 4 weeks of cardiac monitoring. Patients with haemorrhagic stroke or with known cause of ischemic stroke, such as known AF or Flutter, symptomatic intracranial/extracranial stenosis (≥50% stenosis) or documented arterial dissections were excluded. Patients with patent foramen ovale (PFO) considered causally related to the index stroke were excluded. PFO-related stroke was defined according to a predefined etiological attribution based on the Risk of Paradoxical Embolism (RoPE) score and the PFO-Associated Stroke Causal Likelihood (PASCAL) classification, and these patients were therefore classified as having a non-ESUS stroke etiology^1^.

### Study variables and data management

The dataset included codified parameters regarding demographic data, vascular risk factors, blood biomarkers, imaging data, cardiac work up and stroke severity rated by the National Institute of Health Stroke Scale (NIHSS)

The cardiac workup comprised left atrial enlargement, complex aortic arch plaques or mobile thrombus, left ventricular dysfunction, mitral valve stenosis, and AFDAS. Left atrial enlargement was assessed by TTE using indexed left atrial area (cm²), in accordance with ASE/EACVI recommendations^18^. Complex aortic arch plaques were identified by TEE and defined according to standard criteria as plaques ≥4 mm in thickness, plaques with ulceration, or plaques with mobile components^45^. Left ventricular dysfunction was defined as the presence of regional wall motion akinesia or a left ventricular ejection fraction <30%, calculated using the biplane Simpson’s method.

Mitral valve stenosis was initially recorded as part of the cardiac evaluation and assessed by TTE; however, patients in whom mitral stenosis was considered a likely cause of stroke were excluded from the study. Due to its very low prevalence in the remaining cohort, mitral valve stenosis was not included as a predictive target in the machine learning models.

Continuous cardiac rhythm monitoring was used to identify AFDAS, defined as episodes lasting more than 30 seconds. AFDAS was collected for descriptive purposes only and was not included as a predictor in any of the machine learning models, as it represents a post-diagnostic finding not available at the time of ESUS evaluation.

Potential competing stroke etiologies were defined as the presence of more than one plausible cardio-aortic embolic source identified during the diagnostic workup. These cases, accounting for approximately 8% of the study population, were retained to reflect real-world diagnostic complexity, in which etiological attribution is often probabilistic rather than binary.

All selected data was entered into an electronic database specifically designed for the study, ensuring patient anonymity and compliance with data privacy and personal data protection regulations (EU Regulation 2016/679).

### Outcome definitions

The predictive task targeted three potential cardio-aortic embolic sources: left atrial enlargement (LAE), left ventricular akinesia or dysfunction (LVD), and complex aortic plaque (AP). Each outcome was modelled independently, using binary classification (presence = 1, absence = 0) as the dependent variable. Potential competing etiologies were defined as the presence of more than one plausible cardio-aortic embolic source identified during the diagnostic workup. In these cases, each embolic source was modeled independently, allowing a single patient to contribute as a positive case to more than one target. This approach reflects the probabilistic nature of etiological attribution in ESUS and avoids forced assignment to a single dominant mechanism.

### Sample size calculation

No formal a priori sample size calculation was performed due to the retrospective design of the study. Sample size adequacy and model robustness were evaluated through a series of sensitivity analyses, including learning curve analyses and bootstrap-based assessments of discrimination, calibration, and feature importance stability (1,000 resamples). These analyses were designed to empirically assess whether model performance and key predictors were stable across different data perturbations. Full methodological details are provided in the Supplementary Methods.

### Preliminary Statistical Analysis

A descriptive statistical analysis was performed to characterize the study variables. Continuous variables were expressed as means ± standard deviation or median with interquartile range, depending on data distribution. Normal distribution was assessed by histogram Normal curve overlay and Kolmogorov-Smirnov Test. Categorical variables were presented as absolute numbers and percentages. An analysis of associations between clinical, laboratory, and imaging variables and echocardiographic targets was conducted to perform an initial feature selection for inclusion in the ML model. The association between clinical variables and echocardiographic targets was preliminarily assessed using correlation tests (Pearson), chi-square, and Mann-Whitney U tests as appropriated. The analysis was performed using R (version 4.3.1) within the RStudio environment (version 2024.12.1).

### Model development and feature interpretation

Three supervised learning approaches—XGBoost (XGB), Random Forest (RF), and a regularized logistic regression model (LR) as a classical benchmark—were evaluated to predict the presence of three potential cardio-aortic embolic sources in ESUS: left atrial enlargement (LAE), left ventricular dysfunction or akinesia (LVD), and complex aortic plaque (AP). Each outcome was modeled independently using the same methodological framework to ensure comparability across targets.

The dataset was split into a training set (80%) and an independent hold-out test set (20%), stratified by outcome. The test set was reserved exclusively for final evaluation and uncertainty estimation and was not used for feature selection, hyperparameter tuning, or threshold selection. Model development was performed within the training set using stratified 10-fold cross-validation.

Preprocessing was implemented within a unified scikit-learn pipeline to ensure that all transformations were fit only on training data within each cross-validation fold.

Continuous variables were imputed using multivariate iterative imputation and categorical variables using mode imputation. Continuous predictors were standardized (z-score), and categorical predictors were one-hot encoded. No random under-sampling or over-sampling techniques were applied. Class imbalance was handled at the algorithm level through class weighting when supported (e.g., optimization of XGBoost scale_pos_weight).

Hyperparameters were optimized using Bayesian optimization (Optuna) within cross-validation, with overfitting controlled through regularization and early stopping. Final performance was assessed on the independent hold-out test set using threshold-free and threshold-dependent metrics, including ROC-AUC, PR-AUC, accuracy, balanced accuracy, precision, recall, and F1-score. Calibration was evaluated using the Brier score and calibration curves. Uncertainty in test-set performance was quantified using bootstrap resampling.

Comparative performance across models was assessed using the Wilcoxon signed-rank test, with PR-AUC as the primary comparison metric due to its suitability for imbalanced outcomes^19^. In cases where the PR-AUC comparison did not reveal clear differences, the F1 scores were examined due to its efficacy in showing model’s overall performances and in handling both false positives and false negatives^20^.

Model interpretability was assessed using permutation importance and SHAP (SHapley Additive exPlanations). Permutation importance quantifies the decrease in model performance observed when the values of a given variable are randomly permuted, thereby providing a global ranking of each feature’s contribution to predictive accuracy^21^. SHAP analyses decompose individual model predictions into additive feature contributions, allowing characterization of the direction, shape, and non-linear patterns of feature effects across the population^22^. To avoid misinterpretation of model-derived quantities as causal or effect-size estimates, SHAP values were interpreted qualitatively and were not used to derive quantitative effect measures. Given that XGBoost achieved the best overall performance in internal validation, subsequent analyses focused on this model, while RF and LR results are reported as benchmark comparators. Full implementation details, hyperparameter ranges, and sensitivity analyses are provided in the Supplementary Materials.

### Ethics and data governance

All data were de-identified and stored in a secure institutional database in compliance with the European Union General Data Protection Regulation (GDPR 2016/679). Given the retrospective nature of the study and the use of fully anonymized registry data, the requirement for informed consent was waived by the institutional ethics committee.

## Results

### Study population

Among 9,299 patients with ischemic stroke recorded in the Vall d’Hebron subdataset of the Catalan Stroke Registry, 2,231 met diagnostic criteria for ESUS. After application of exclusion criteria, 1,741 patients were included in the final analysis (Figure 1).

**Figure 1.**
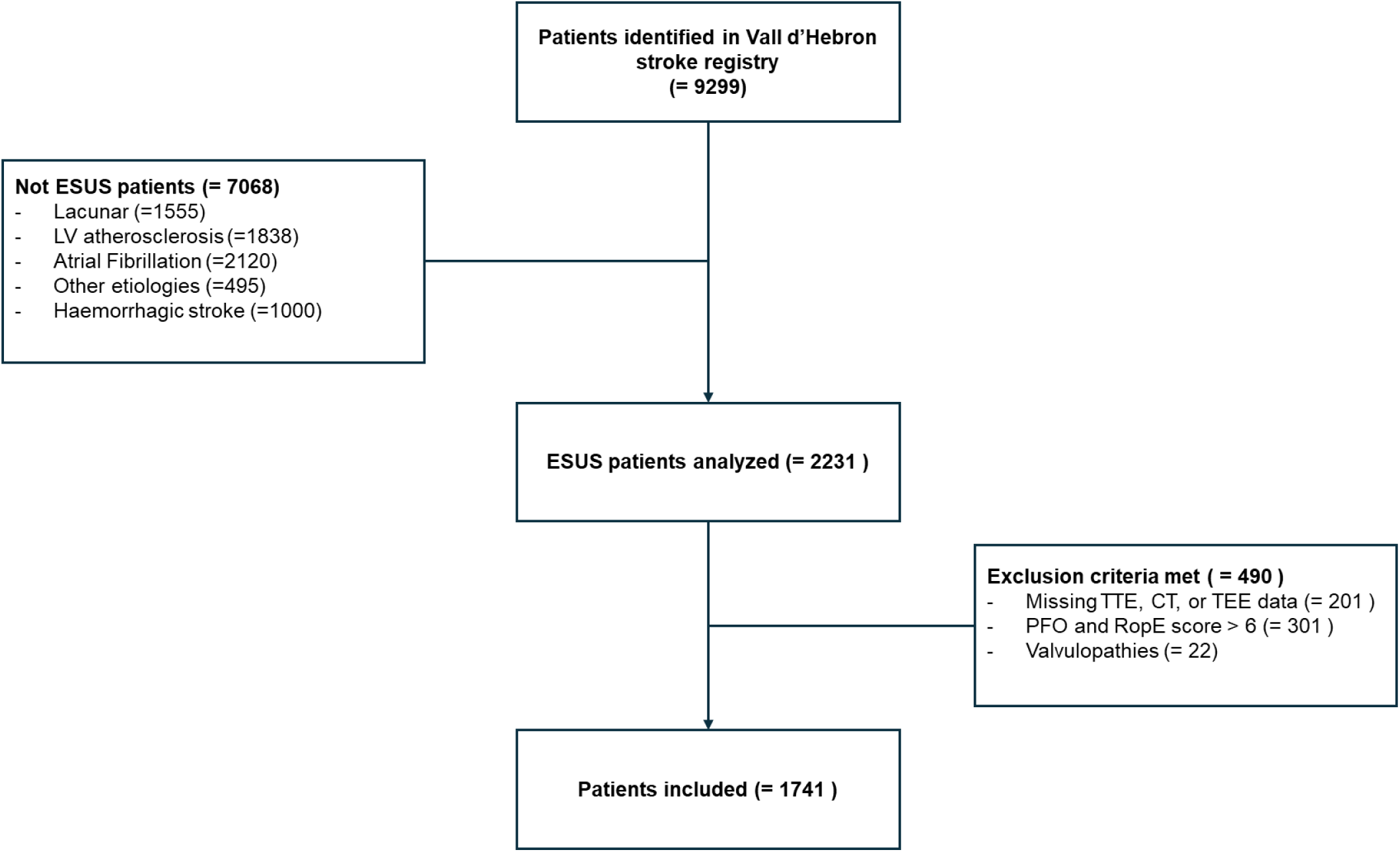
**Patients inclusion flowchart**. Legend: ESUS: Embolic Stroke of Undetermined Source, LV: large vessel, TTE: transthoracic echocardiography, CT: computed tomography, TEE: transesophageal echocardiography, PFO: patent foramen ovale, ROPE: Risk of Paradoxical Embolism.

The mean age was 71.5 ± 14.6 years, and 48.3% were women. The most prevalent vascular risk factors were hypertension (59.0%), dyslipidemia (41.5%), type 2 diabetes (24.3%), and smoking (13.0%). Ischemic heart disease was present in 8.3% of patients and chronic kidney disease in 2.2%. A previous transient ischemic attack or stroke was reported in 8.2%, and 4.1% experienced stroke recurrence within one year.

At admission, the median NIHSS score was 3 (IQR 1–9). Vessel occlusion was identified in 24.6% of cases, whereas 75.4%—including 20.3% transient ischemic attacks—showed no arterial occlusion. The median NT-proBNP level was 378 pg/mL (IQR 127–1,284).

Regarding potential embolic sources, left atrial enlargement was identified in 40.5% of patients, complex aortic plaques in 11.0%, and left ventricular dysfunction in 6.5%.

Atrial fibrillation was detected in 9.6% of patients, with 6.3% identified during inpatient telemetry and an additional 3.3% detected through one-month post-discharge Holter monitoring.

Overlapping embolic sources were uncommon. Specifically, 3.6% of patients had both LAE and LVD, 3.3% had LAE and AP, 1.7% had AP and LVD, and only 0.3% presented with all three conditions (Table 1).

**Table 1.**
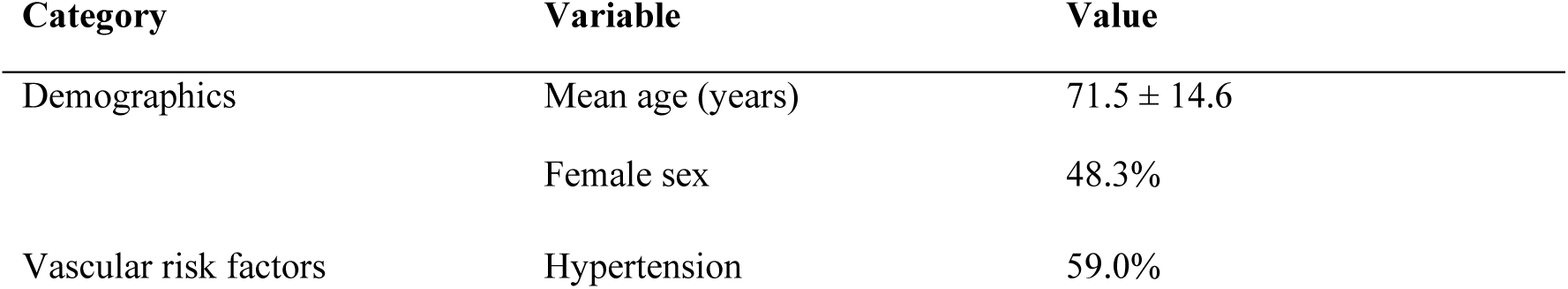
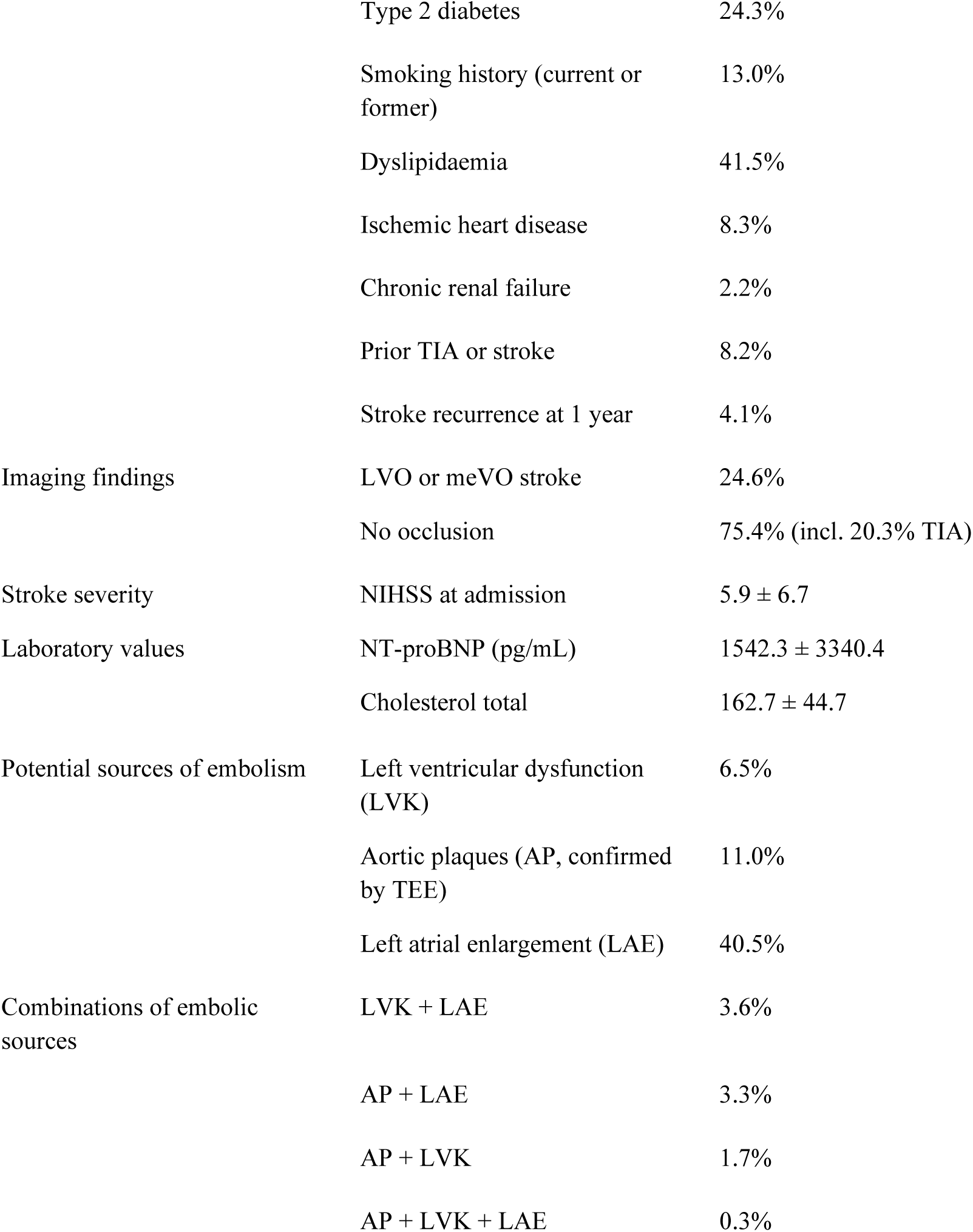
Baseline demographic, clinical, and imaging characteristics of patients with embolic stroke of undetermined source (n = 1,741).

### Influence of the Variables in Each Model

Across the three targets, XG Boost consistently provided the best overall performance that are listed in Table 2.

**Table 2.** XGBoost performances among the three targets.

| Model | PR-AUC | Balanced Accuracy | Sensitivity | Specificity | F1 score (class 1) | Precision |
| --- | --- | --- | --- | --- | --- | --- |
| LAE | 0.71 | 0.71 | 0.67 | 0.67 | 0.70 | 0.77 (Class 0)<br>0.65 (Class 1) |
| AP | 0.29 | 0.58 | 0.79 | 0.54 | 0.40 | 0.92 (class 0)<br>0.40 (class 1) |
| LVD | 0.44 | 0.76 | 0.54 | 0.96 | 0.50 | 0.97 (class 0)<br>0.47 (class 1) |

Out of 73 baseline variables, the ESUS AI algorithm identified a restricted set of features with consistent relevance across the three predictive models, highlighting distinct yet partially overlapping etiological profiles (Figure 2). Age, NT-proBNP levels, NIHSS emerged as common contributors, although their relative importance and direction of association differed across embolic mechanisms (Figure 2 – 5)

**Figure 2.**
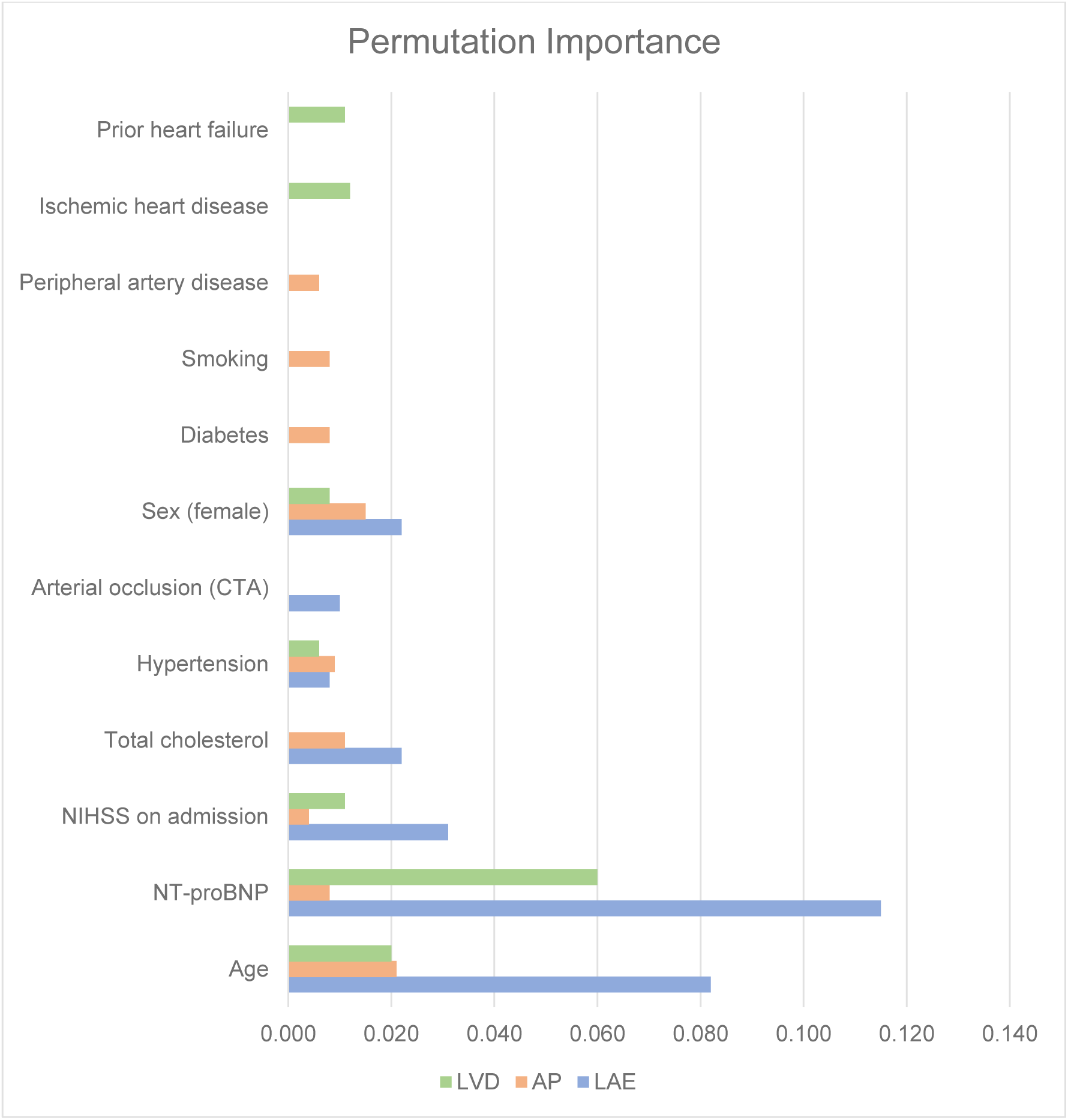
**Permutation Importance analysis**. Age, NT-proBNP, NIHSS in and Cholesterol level represent the most important features for all the three models. Legend: LAE: left atrial enlargement; AP: aortic plaques; LVD: left ventricle dysfunction

### Left atrial enlargement model

In the LAE model, age (PI = 0.082) and NT-proBNP (PI = 0.121) were the dominant contributors to model performance, followed by NIHSS on admission (PI = 0.031). SHAP analyses showed a largely monotonic increase in LAE probability with advancing age and increasing NT-proBNP levels (Figure 2 - 3A). The association between NT-proBNP and LAE was particularly pronounced in older patients, with a nearly linear increase in predicted probability among individuals aged over 80 years (Figure 4A).

**Figure 3.**
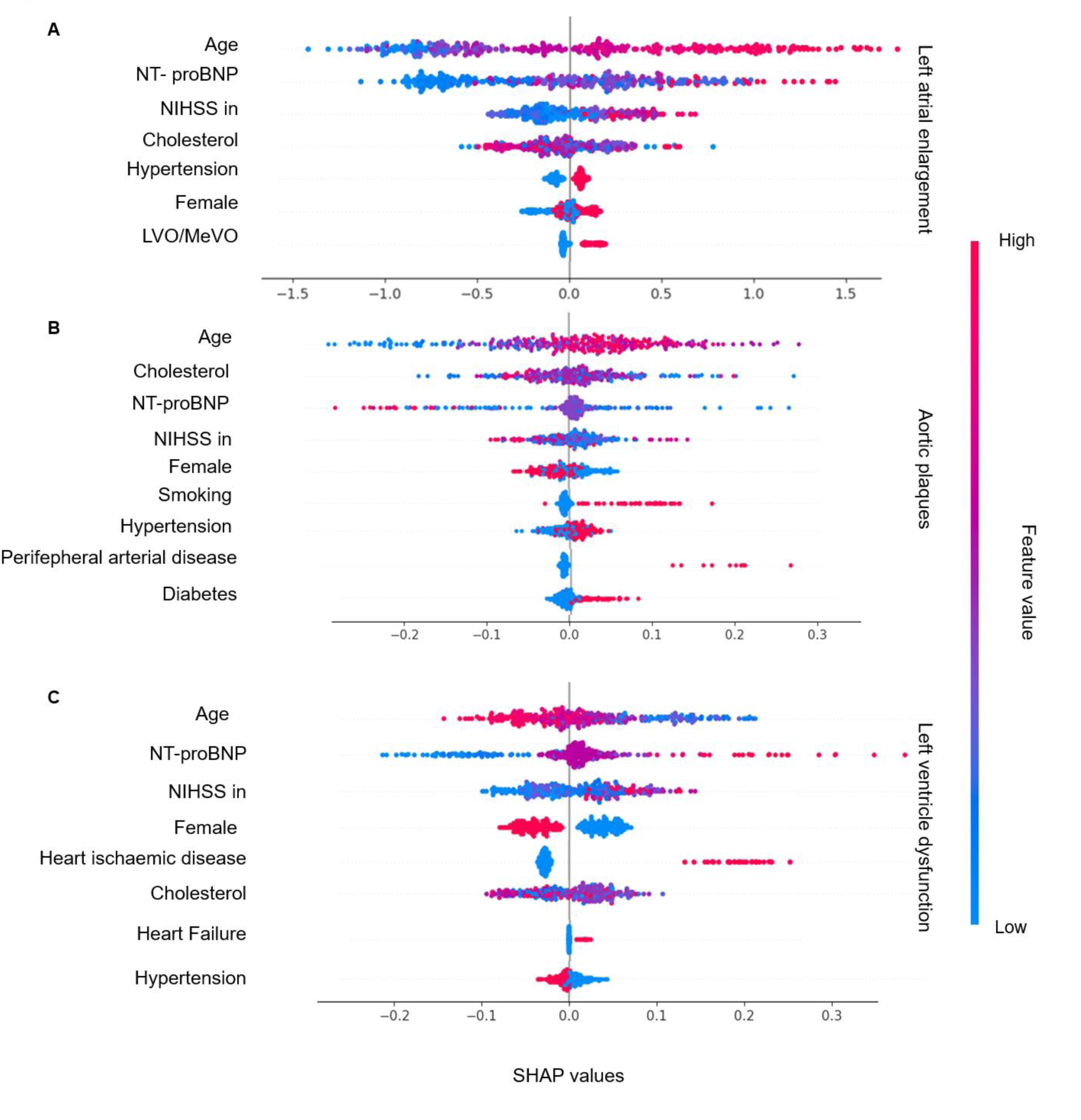
SHAP summary plot of the three models. The x-axis represents the impact of each feature on model predictions, with points to the left indicating a lower probability of target presence (negative values) and points to the right indicating a higher probability of target presence (positive values). The color scale denotes feature value intensity (red: higher values, blue: lower values). A) Higher values of age, NT-proBNP, NIHSS on admission, hypertension, and female sex were associated with an increased likelihood of Left atrial enlargement (red dots in positive SHAP values). B) Age, male sex, smoking, hypertension, peripheral arterial disease, and diabetes demonstrated a positive association with the presence of aortic plaques (red dots in positive SHAP values). C) Lower age, higher NT-proBNP levels, higher NIHSS scores on admission, male sex, and the presence of ischemic heart disease, and heart failure were positively associated with LVD. Cholesterol exhibited an unclear, non-linear effect, while hypertension showed an inverse relationship with left ventricle akinesia probability.

**Figure 4.**
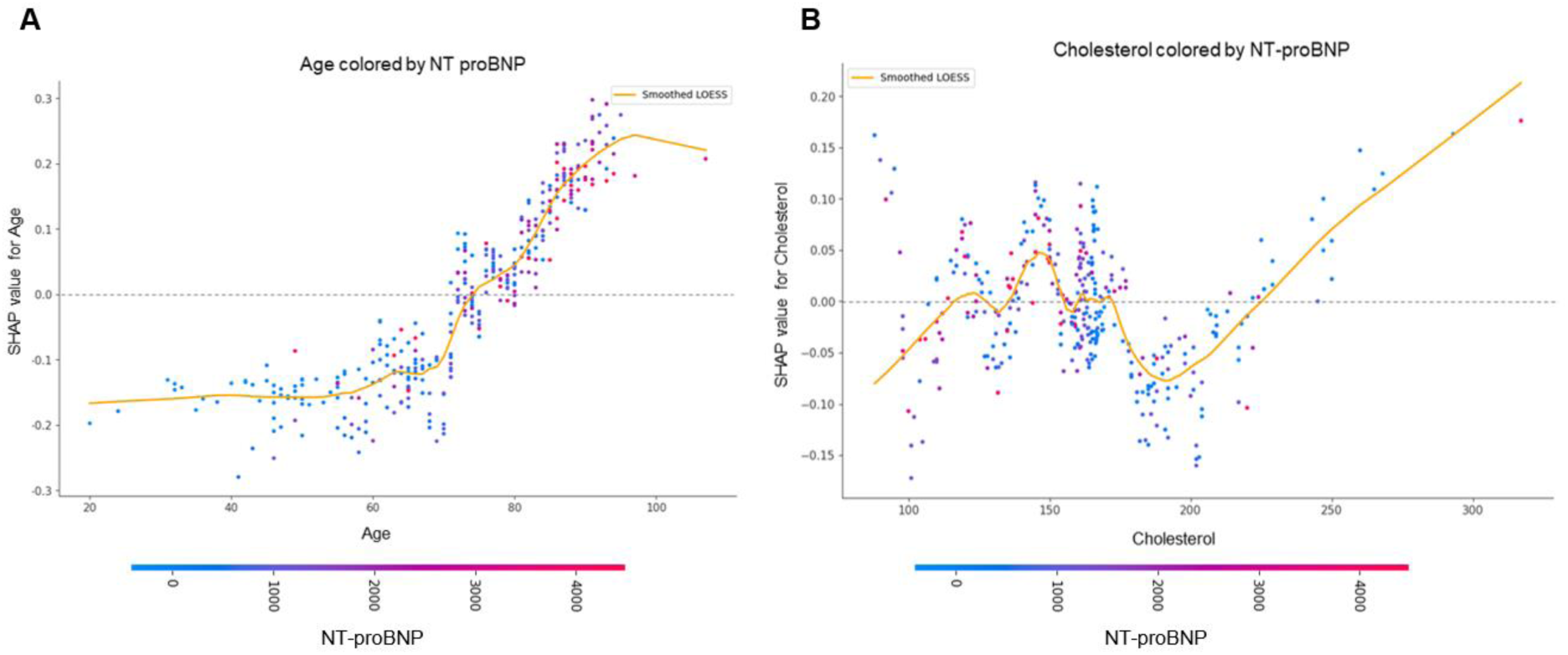
**Left atrial enlargement SHAP dependency plots**. In these plots, the x-axis shows the values of the first feature, the y-axis indicates the predicted probability of LAE, with point above the dotted line representing high probability of LAE presence, and below high probability of LAE absence. The color of each data point reflects the value of a secondary interacting feature. A): Age and NT-proBNP showed approximately linear relationships with LAE probability, each amplifying the effects of the others. B): Cholesterol exhibited a distinctly non-linear relationship: very low in patients with high NT-proBNP, and very high cholesterol values were associated with increased LAE risk, whereas normal cholesterol levels correlated with decreased LAE probability.

**Figure 5.**
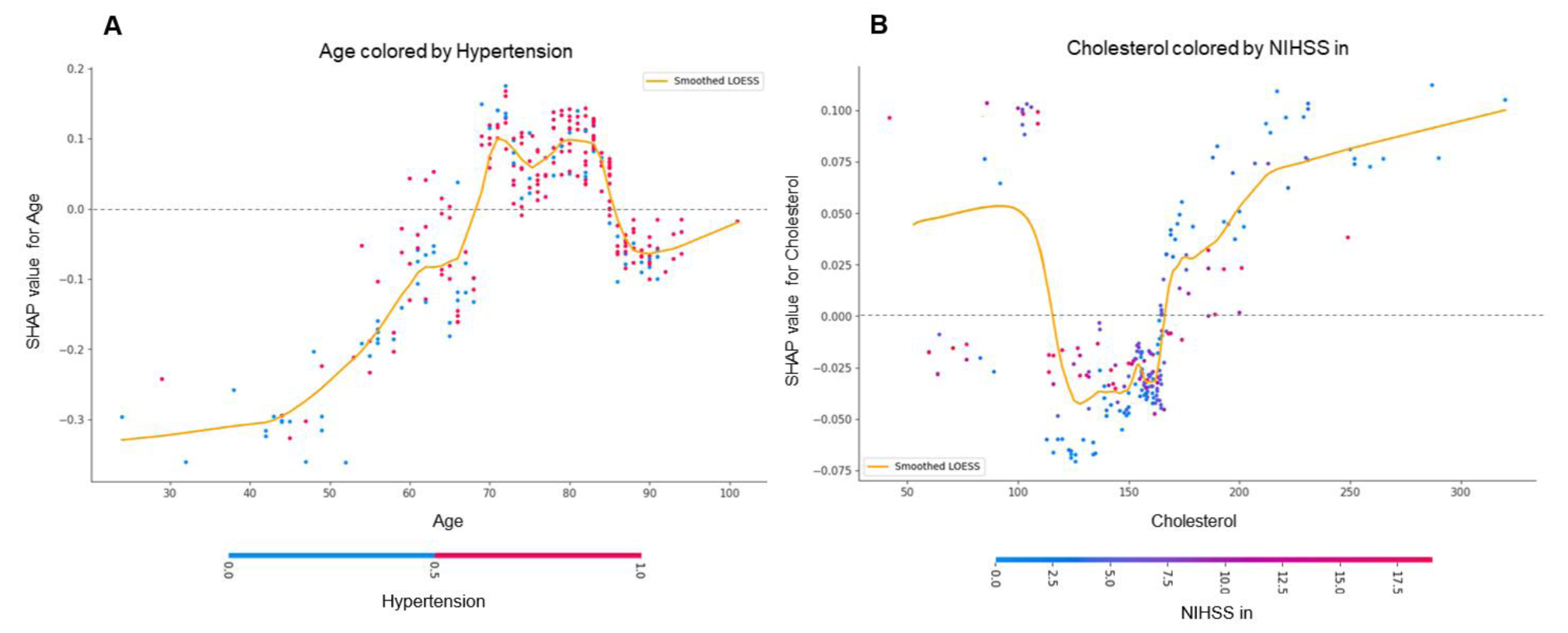
Aortic plaques SHAP dependency plots. A) Older age consistently correlated with an increased probability of AP, in particular this effect is amplified by hypertension. Cholesterol exhibited greater dispersion of predictions at low values (<100 mg/dL); however, for values above 100 mg/dL, the relationship became more linear, with higher cholesterol levels increasingly predicting the presence of AP, without clear correlation with NIHSS.

Total cholesterol displayed a non-linear relationship with LAE (PI = 0.022), with both low (<150 mg/dL) and very high (>250 mg/dL) values associated with increased LAE probability (Figure 4B). Additional contributors included hypertension (PI = 0.008), presence of arterial occlusion on CTA (PI = 0.010), and sex (0.018), each exerting a smaller but consistent influence on model predictions (Figure 2 – 3A).

### Aortic Plaque Model

The AP model was primarily driven by age and traditional vascular risk factors. Permutation importance ranked age as the most influential feature (PI = 0.021), followed by hypertension (PI = 0.011), cholesterol levels (PI = 0.011), and other atherosclerotic risk factors such as diabetes (PI = 0.008), smoking (PI = 0.008), and peripheral artery disease (PI = 0.006) (Figure 2 - 3B).

SHAP dependency plots demonstrated a progressive increase in AP probability with advancing age, which was amplified in hypertensive patients and at higher cholesterol levels (Figures 5A–B). In contrast, NT-proBNP levels and NIHSS showed an inverse association with AP probability, suggesting that patients with more severe strokes or higher cardiac biomarker levels were less likely to harbor complex aortic plaques as the dominant embolic source.

### Left Ventricular Dysfunction

In the LVD model, NT-proBNP emerged as the single most influential predictor (PI = 0.060), followed by age (PI = 0.020) and NIHSS (PI = 0.011) (Figure 2 – 3C). SHAP analyses revealed that elevated NT-proBNP levels were strongly associated with increased LVD probability, particularly at values exceeding 2000 pg/mL (Figure 6).

**Figure 6.**
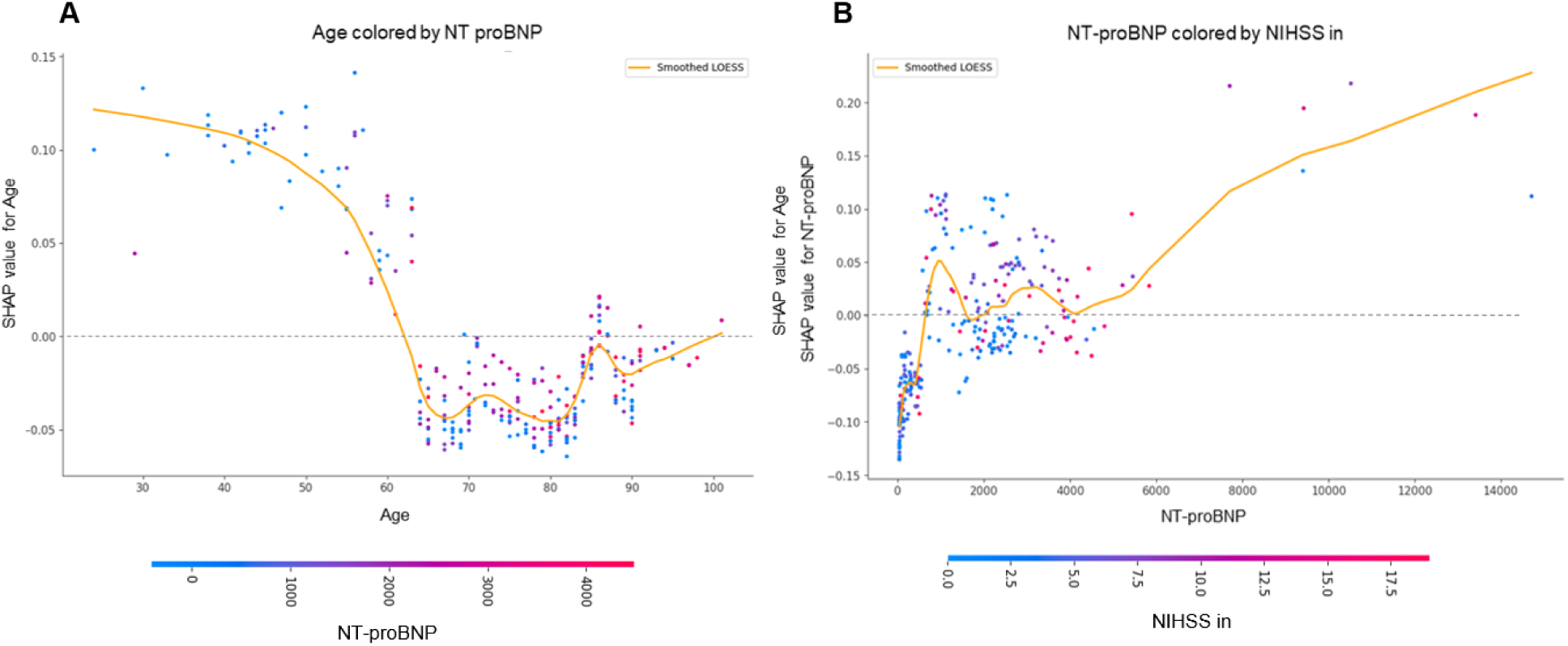
Left ventricle dysfunction SHAP dependency plots. A) B) Lower age and higher NT-proBNP values consistently increased the likelihood of LVD.

Interestingly, younger age was associated with a higher likelihood of LVD, indicating a distinct etiological pattern compared with LAE and AP. A history of ischemic heart disease (PI = 0.012), and prior heart failure (PI = 0.011) further increased LVD probability, while male sex (PI = 0.008) contributed modestly to model predictions (Figure 2 – 3C).

### Clinical case

To illustrate how the model can inform clinical decisions, consider a fictitious 77-year-old male with ESUS due to a right M2 occlusion, NIHSS 14, hypertension, dyslipidemia, no cardiac disease, NT-proBNP 958 pg/mL, and cholesterol 235 mg/dL.

For this profile, the model yields approximate probabilities of 0.78 for LAE, 0.20 for AP, and 0.07 for LVD. In this pattern algorithm would support a diagnostic strategy prioritizing prolonged rhythm monitoring (e.g., external or implantable loop recorder) over extensive aortic imaging or left-ventricle-focused investigations (see Figure 7)

**Figure 7.**
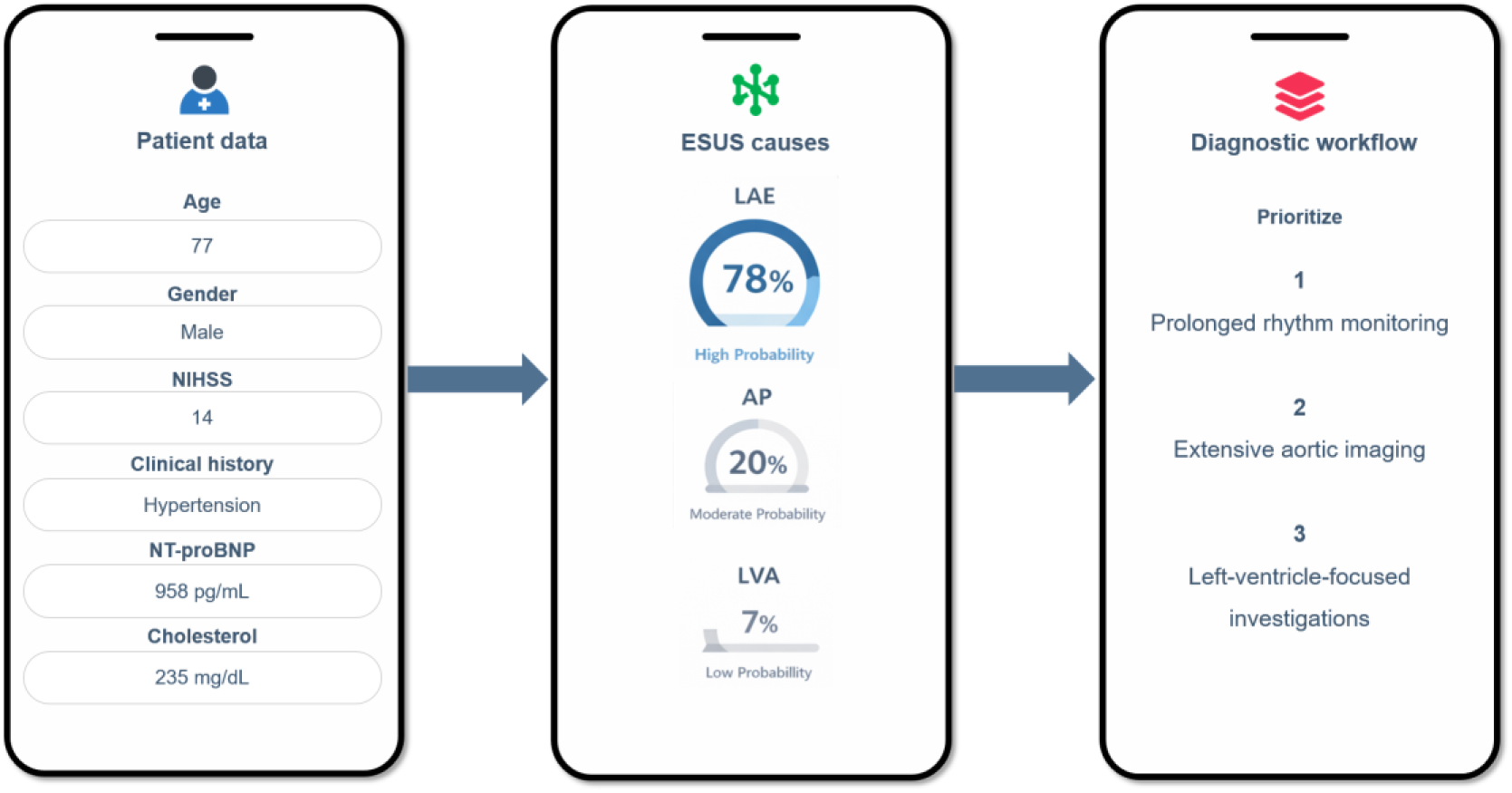
**Conceptual example of a ESUS AI–based clinical decision support application**. Patient clinical and laboratory data are entered into the calculator (left panel), which generates individualized probability estimates for the main suspected ESUS pathophysiological mechanisms—left atrial enlargement (LAE), aortic plaques (AP), and left ventricular akinesia (LVD) (center panel). These probabilities are then translated into a prioritized, patient-specific diagnostic workflow (right panel), supporting clinicians in tailoring cardiac and vascular investigations. Original creation using PowerPoint

## Discussion

ESUS represents a diagnostic phase in which the underlying embolic source remains unproven despite extensive standard workup. Although each ischemic stroke arises from a single embolic mechanism, the frequent inability to identify that mechanism in ESUS has limited the effectiveness of uniform secondary prevention strategies stratification^3–5^.

Using supervised machine-learning methods in a real-world ESUS cohort undergoing standardized diagnostic evaluation, we identified three reproducible and biologically coherent etiologic profiles—left atrial enlargement, complex aortic plaques, and left ventricular dysfunction—each characterized by distinct combinations of demographic features, vascular risk factors, stroke severity, and cardiac biomarkers.

Rather than assigning a deterministic etiologic diagnosis, our approach estimates the relative probability of candidate embolic sources, with the explicit aim of orienting and prioritizing the post-stroke diagnostic workflow. In ESUS, establishing the true embolic mechanism often requires extensive, time-consuming, and costly investigations that cannot realistically be applied indiscriminately to all patients. By leveraging routinely collected data, this framework supports a personalized, hypothesis-driven diagnostic strategy, improving efficiency while fully preserving established etiologic principles.

### LAE risk profile

Patients classified within the LAE profile were typically older, had elevated NT-proBNP levels, and presented with more severe strokes, frequently associated with large-vessel occlusions. Although LAE is not a direct embolic mechanism, it is a well-established marker of atrial cardiomyopathy and occult atrial fibrillation^25–28^. Its identification is clinically relevant, as it can guide intensified and prolonged rhythm monitoring strategies aimed at improving AF detection.

Age showed a nearly monotonic association with LAE probability, consistent with progressive atrial remodeling and fibrosis observed with aging^29,30^. NT-proBNP emerged as a dominant contributor, reflecting atrial stretch and combined atrial–ventricular dysfunction^31^ ^32–34^. Notably, the association between NT-proBNP and LAE was non-linear, with a tendency toward saturation at very high values, which may reflect the contribution of non-cardiac conditions such as renal dysfunction, systemic inflammation, or hypoxic states^35,36^.

Total cholesterol demonstrated a non-linear relationship with LAE, with increased probability observed at both low and very high levels. This finding is consistent with the so-called cholesterol paradox previously described in atrial pathology and may reflect complex interactions between lipid metabolism, systemic inflammation, and atrial structural remodeling^37^ ^38,39^. LAE risk also rose at very high cholesterol levels (>250 mg/dL), likely reflecting clustering with other cardiovascular risk factors ^42^.

Stroke severity, as reflected by NIHSS, also contributed to the LAE profile, likely acting as a marker of embolic burden due to higher severity and mortality of cardioembolic strokes^43,44^.

### AP risk profile

The second profile identified by the model corresponded to a predominantly atherosclerotic pattern, characterized by older age, male sex, and a high burden of traditional vascular risk factors, including hypertension, diabetes, smoking, and elevated cholesterol levels.

These findings are consistent with prior evidence indicating that complex aortic plaques, particularly those ≥4 mm or with ulcerated or mobile components, represent a significant but often under-recognized embolic source^45^. The inverse association observed between stroke severity and AP probability may reflect the chronic nature of atherosclerotic disease, with plaque rupture leading to multiple distal microemboli rather than large proximal occlusions^51^. Additionally, the presence of long-standing vascular disease may promote collateral circulation, contributing to milder clinical presentations^48–50^.

### LVD risk profile

The third profile was defined by a cardiomyopathic pattern, characterized by elevated NT-proBNP levels, higher stroke severity, male sex, and a history of cardiac disease, particularly ischemic cardiomyopathy. Interestingly, younger age was associated with a higher probability of LVD, distinguishing this profile from LAE and AP.

While ischemic heart disease remains the most common cause of left ventricular dysfunction^52^, non-ischemic cardiomyopathies (such as dilated or non-compaction cardiomyopathy, drug toxicity, and Takotsubo syndrome) - more prevalent in younger individuals—can also lead to regional wall motion abnormalities and intraventricular blood stasis^53^. This constellation of features delineates a mechanism in which ventricular akinesia and impaired systolic function increase the risk of ventricular thrombus formation, leading to severe cardioembolic strokes^55,56^. As observed in the WATCH trial and other studies, reduced ejection fraction and ventricular dilation are key determinants of thromboembolic risk in this population^54^.

In this context, the inclusion of additional biomarkers such as cardiac troponins could have further refined pathophysiological discrimination between acute myocardial injury and chronic ventricular dysfunction. However, troponin levels were not routinely available in the ESUS diagnostic workup at our institution and were therefore not included in the present analysis.

### Comparison of the performances of the models pattern among the targets and clinical usefulness

The LAE model demonstrated balanced performance in identifying both the presence and absence of left atrial enlargement, supporting its robustness for etiological stratification in ESUS. In contrast, the AP and LVD models showed asymmetric performance patterns, likely reflecting the lower prevalence and greater heterogeneity of these conditions. Despite this, both models retain clinical value when interpreted within a targeted diagnostic framework. The AP model favored sensitivity, making it suitable as a screening tool to prioritize patients for transesophageal echocardiography and minimize missed aortic sources. Conversely, the LVD model achieved high specificity and negative predictive value, supporting its use in ruling out ventricular dysfunction and potentially reducing unnecessary advanced cardiac imaging in patients with low predicted risk.

### Strengths, limitations, and future perspectives

The proposed ESUS AI framework may support a more efficient post-stroke diagnostic workup by helping clinicians prioritize targeted investigations and streamline cardiac diagnostic pathways. This approach may be particularly useful for non-expert neurologists and for centers with limited access to advanced diagnostic resources, where structured guidance can optimize the allocation of cardiological investigations.

Importantly, the model was designed by clinicians to address real-world unmet needs and was intentionally developed as a transparent and interpretable system, combining a high-performing machine learning architecture with post-hoc explainability through SHAP analyses to mitigate black-box behavior and promote reproducibility.

Several limitations should be acknowledged. First, atrial fibrillation was not included as a direct target outcome; however, left atrial enlargement was used as an imaging-based surrogate of atrial cardiomyopathy, which is mechanistically linked to occult AF. Second, the single-center design may limit generalizability, although the standardized diagnostic protocol enhances internal validity. Third, the lack of external validation remains a key limitation and precludes immediate clinical implementation. In addition, some predictors such as stroke severity may reflect embolic burden rather than causality, potentially introducing circularity.

Future directions include external multicenter validation and the integration of additional clinical, laboratory, and imaging biomarkers to further refine etiological stratification. Collaborative development within shared research platforms could facilitate independent validation, continuous model refinement, and broader clinical applicability.

## Data Availability

The data will be shared upon request

## 6. Acknowledgments

We thank the Stroke Unit and Cardiology teams at Hospital Universitari Vall d’Hebron, including nurses and physicians, for their essential contribution to patient care and diagnostic workup.

## References

1. Ntaios G, Baumgartner H, Doehner W, Donal E, Edvardsen T, Healey JS, Iung B, Kamel H, Kasner SE, Korompoki E, et al. Embolic strokes of undetermined source: a clinical consensus statement of the ESC Council on Stroke, the European Association of Cardiovascular Imaging and the European Heart Rhythm Association of the ESC. European Heart Journal. 2024;45:1701–1715.

2. Hart RG, Diener H-C, Coutts SB, Easton JD, Granger CB, O’Donnell MJ, Sacco RL, Connolly SJ. Embolic strokes of undetermined source: the case for a new clinical construct. The Lancet Neurology. 2014;13:429–438.

3. Kamel H, Longstreth WT Jr, Tirschwell DL, Kronmal RA, Marshall RS, Broderick JP, Aragón García R, Plummer P, Sabagha N, Pauls Q, et al. Apixaban to Prevent Recurrence After Cryptogenic Stroke in Patients With Atrial Cardiopathy: The ARCADIA Randomized Clinical Trial. JAMA. 2024;331:573–581.

4. Diener H-C, Sacco RL, Easton JD, Granger CB, Bernstein RA, Uchiyama S, Kreuzer J, Cronin L, Cotton D, Grauer C, et al. Dabigatran for Prevention of Stroke after Embolic Stroke of Undetermined Source. New England Journal of Medicine. 2019;380:1906–1917.

5. Hart RG, Sharma M, Mundl H, Kasner SE, Bangdiwala SI, Berkowitz SD, Swaminathan B, Lavados P, Wang Y, Wang Y, et al. Rivaroxaban for Stroke Prevention after Embolic Stroke of Undetermined Source. New England Journal of Medicine. 2018;378:2191–2201.

6. Tullio MRD, Homma S, Jin Z, Sacco RL. Aortic Atherosclerosis, Hypercoagulability and Stroke: the Aortic Plaque and Risk of Ischemic Stroke (APRIS) Study. Journal of the American College of Cardiology. 2008;52:855.

7. Yoshida Y, Jin Z, Mannina C, Homma S, Nakanishi K, Leibowitz D, Elkind MSV, Rundek T, Di Tullio MR. Aortic Arch Plaques and the Long-Term Risk of Stroke and Cardiovascular Events in the Statin Era. Stroke. 2024;55:69–77.

8. Rubiera M, Aires A, Antonenko K, Lémeret S, Nolte CH, Putaala J, Schnabel RB, Tuladhar AM, Werring DJ, Zeraatkar D, et al. European Stroke Organisation (ESO) guideline on screening for subclinical atrial fibrillation after stroke or transient ischaemic attack of undetermined origin. European Stroke Journal. 2022;7:CVII–CXXXIX.

9. Caso V, Turc G, Abdul-Rahim AH, Castro P, Hussain S, Lal A, Mattle H, Korompoki E, Søndergaard L, Toni D, et al. European Stroke Organisation (ESO) Guidelines on the diagnosis and management of patent foramen ovale (PFO) after stroke. European Stroke Journal. 2024;23969873241247978.

10. Ning Y, Tse G, Luo G, Li G. Atrial Cardiomyopathy: An Emerging Cause of the Embolic Stroke of Undetermined Source. Front. Cardiovasc. Med. [Internet]. 2021 [cited 2024 Oct 21];8. Available from: https://www.frontiersin.org/journals/cardiovascular-medicine/articles/10.3389/fcvm.2021.674612/full

11. Park H-K, Kim BJ, Yoon C-H, Yang MH, Han M-K, Bae H-J. Left Ventricular Diastolic Dysfunction in Ischemic Stroke: Functional and Vascular Outcomes. J Stroke. 2016;18:195–202.

12. Ramasamy S, Yaghi S, Salehi Omran S, Lerario MP, Devereux R, Okin PM, Gupta A, Navi BB, Kamel H, Merkler AE. Association Between Left Ventricular Ejection Fraction, Wall Motion Abnormality, and Embolic Stroke of Undetermined Source. Journal of the American Heart Association. 2019;8:e011593.

13. Zietz A, Polymeris AA, Helfenstein F, Schaedelin S, Hert L, Wagner B, Seiffge DJ, Traenka C, Altersberger VL, Dittrich T, et al. The impact of competing stroke etiologies in patients with atrial fibrillation. Eur Stroke J. 2023;8:703–711.

14. McComb M, Blair RH, Lysy M, Ramanathan M. Machine learning-guided, big data-enabled, biomarker-based systems pharmacology: modeling the stochasticity of natural history and disease progression. J Pharmacokinet Pharmacodyn. 2022;49:65–79.

15. Weissler EH, Naumann T, Andersson T, Ranganath R, Elemento O, Luo Y, Freitag DF, Benoit J, Hughes MC, Khan F, et al. The role of machine learning in clinical research: transforming the future of evidence generation. Trials. 2021;22:537.

16. Intel·ligència analítica [Internet]. Agència de Qualitat i Avaluació Sanitàries de Catalunya (AQuAS). [cited 2025 May 5];Available from: http://aquas.gencat.cat/ca/fem/intelligencia-analitica

17. Checklist for Artificial Intelligence in Medical Imaging (CLAIM) [Internet]. Radiology: Artificial Intelligence. [cited 2025 Dec 9];Available from: https://pubs.rsna.org/page/ai/claim?doi=10.1148%2Fryai&publicationCode=ai

18. Nagueh SF, Smiseth OA, Appleton CP, Byrd BF, Dokainish H, Edvardsen T, Flachskampf FA, Gillebert TC, Klein AL, Lancellotti P, et al. Recommendations for the Evaluation of Left Ventricular Diastolic Function by Echocardiography: An Update from the American Society of Echocardiography and the European Association of Cardiovascular Imaging. J Am Soc Echocardiogr. 2016;29:277–314.

19. Saito T, Rehmsmeier M. The precision-recall plot is more informative than the ROC plot when evaluating binary classifiers on imbalanced datasets. PLoS One. 2015;10:e0118432.

20. Owusu-Adjei M, Hayfron-Acquah JB, Frimpong T, Abdul-Salaam G. Imbalanced class distribution and performance evaluation metrics: A systematic review of prediction accuracy for determining model performance in healthcare systems. PLOS Digital Health. 2023;2:e0000290.

21. Altmann A, Toloşi L, Sander O, Lengauer T. Permutation importance: a corrected feature importance measure. Bioinformatics. 2010;26:1340–1347.

22. Lundberg SM, Lee S-I. A unified approach to interpreting model predictions [Internet]. In: Proceedings of the 31st International Conference on Neural Information Processing Systems. Red Hook, NY, USA: Curran Associates Inc.; 2017 [cited 2026 Jan 31]. p. 4768–4777.Available from: https://dl.acm.org/doi/10.5555/3295222.3295230

23. Kamel H, Navi BB, Parikh NS, Merkler AE, Okin PM, Devereux RB, Weinsaft JW, Kim J, Cheung JW, Kim LK, et al. Machine Learning Prediction of Stroke Mechanism in Embolic Strokes of Undetermined Source. Stroke. 2020;51:e203–e210.

24. Speiser JL, Kerr WT, Ziegler A. Common Critiques and Recommendations for Studies in Neurology Using Machine Learning Methods. Neurology. 2024;103:e209861.

25. Sanfilippo AJ, Abascal VM, Sheehan M, Oertel LB, Harrigan P, Hughes RA, Weyman AE. Atrial enlargement as a consequence of atrial fibrillation. A prospective echocardiographic study. Circulation. 1990;82:792–797.

26. Zacà V, Galderisi M, Mondillo S, Focardi M, Ballo P, Guerrini F. Left atrial enlargement as a predictor of recurrences in lone paroxysmal atrial fibrillation. Can J Cardiol. 2007;23:869–872.

27. Abhayaratna WP, Seward JB, Appleton CP, Douglas PS, Oh JK, Tajik AJ, Tsang TSM. Left Atrial Size: Physiologic Determinants and Clinical Applications. Journal of the American College of Cardiology. 2006;47:2357–2363.

28. Jordan K, Yaghi S, Poppas A, Chang AD, Mac Grory B, Cutting S, Burton T, Jayaraman M, Tsivgoulis G, Sabeh MK, et al. Left Atrial Volume Index Is Associated With Cardioembolic Stroke and Atrial Fibrillation Detection After Embolic Stroke of Undetermined Source. Stroke. 2019;50:1997–2001.

29. Thomas L, Levett K, Boyd A, Leung DYC, Schiller NB, Ross DL. Compensatory changes in atrial volumes with normal aging: is atrial enlargement inevitable? Journal of the American College of Cardiology. 2002;40:1630–1635.

30. Aurigemma GP, Gottdiener JS, Arnold AM, Chinali M, Hill JC, Kitzman D. Left atrial volume and geometry in healthy aging: the Cardiovascular Health Study. Circ Cardiovasc Imaging. 2009;2:282–289.

31. Prastaro M, Paolillo S, Savarese G, Dellegrottaglie S, Scala O, Ruggiero D, Gargiulo P, Marciano C, Parente A, Cecere M, et al. N-terminal pro-b-type natriuretic peptide and left atrial function in patients with congestive heart failure and severely reduced ejection fraction†. European Journal of Echocardiography. 2011;12:506–513.

32. Tsuchida K, Tanabe K. Influence of paroxysmal atrial fibrillation attack on brain natriuretic peptide secretion. J Cardiol. 2004;44:1–11.

33. Seegers J, Zabel M, Grüter T, Ammermann A, Weber-Krüger M, Edelmann F, Gelbrich G, Binder L, Herrmann-Lingen C, Gröschel K, et al. Natriuretic peptides for the detection of paroxysmal atrial fibrillation. Open Heart [Internet]. 2015 [cited 2025 Feb 13];2. Available from: https://openheart.bmj.com/content/2/1/e000182

34. di Biase L, Bonura A, Pecoraro PM, Carbone SP, Di Lazzaro V. Unlocking the Potential of Stroke Blood Biomarkers: Early Diagnosis, Ischemic vs. Haemorrhagic Differentiation and Haemorrhagic Transformation Risk: A Comprehensive Review. Int J Mol Sci. 2023;24:11545.

35. Bar SL, Swiggum E, Straatman L, Ignaszewski A. Nonheart failure-associated elevation of amino terminal pro-brain natriuretic peptide in the setting of sepsis. Can J Cardiol. 2006;22:263–266.

36. Inoue S-I, Murakami Y, Sano K, Katoh H, Shimada T. Atrium as a source of brain natriuretic polypeptide in patients with atrial fibrillation. Journal of Cardiac Failure. 2000;6:92–96.

37. Annoura M, Ogawa M, Kumagai K, Zhang B, Saku K, Arakawa K. Cholesterol paradox in patients with paroxysmal atrial fibrillation. Cardiology. 1999;92:21–27.

38. Lee H, Lee S, Choi E, Han K, Oh S. Low Lipid Levels and High Variability are Associated With the Risk of New-Onset Atrial Fibrillation. Journal of the American Heart Association. 2019;8:e012771.

39. Hanna IR, Heeke B, Bush H, Brosius L, King-Hageman D, Dudley SC, Beshai JF, Langberg JJ. Lipid-lowering drug use is associated with reduced prevalence of atrial fibrillation in patients with left ventricular systolic dysfunction. Heart Rhythm. 2006;3:881–886.

40. Goonasekara CL, Balse E, Hatem S, Steele DF, Fedida D. Cholesterol and cardiac arrhythmias. Expert Rev Cardiovasc Ther. 2010;8:965–979.

41. Guo Y, Lip GYH, Apostolakis S. Inflammation in atrial fibrillation. J Am Coll Cardiol. 2012;60:2263–2270.

42. Krishnan A, Sharma H, Yuan D, Trollope AF, Chilton L. The Role of Epicardial Adipose Tissue in the Development of Atrial Fibrillation, Coronary Artery Disease and Chronic Heart Failure in the Context of Obesity and Type 2 Diabetes Mellitus: A Narrative Review. J Cardiovasc Dev Dis. 2022;9:217.

43. Kamel H, Healey JS. Cardioembolic Stroke. Circ Res. 2017;120:514–526.

44. Lin HJ, Wolf PA, Kelly-Hayes M, Beiser AS, Kase CS, Benjamin EJ, D’Agostino RB. Stroke severity in atrial fibrillation. The Framingham Study. Stroke. 1996;27:1760–1764.

45. Amarenco P, Cohen A, Tzourio C, Bertrand B, Hommel M, Besson G, Chauvel C, Touboul PJ, Bousser MG. Atherosclerotic disease of the aortic arch and the risk of ischemic stroke. N Engl J Med. 1994;331:1474–1479.

46. Jones EF, Kalman JM, Calafiore P, Tonkin AM, Donnan GA. Proximal aortic atheroma. An independent risk factor for cerebral ischemia. Stroke. 1995;26:218–224.

47. Harloff A, Simon J, Brendecke S, Assefa D, Helbing T, Frydrychowicz A, Weber J, Olschewski M, Strecker C, Hennig J, et al. Complex plaques in the proximal descending aorta: an underestimated embolic source of stroke. Stroke. 2010;41:1145–1150.

48. Liebeskind DS, Cotsonis GA, Saver JL, Lynn MJ, Cloft HJ, Chimowitz MI, Warfarin–Aspirin Symptomatic Intracranial Disease (WASID) Investigators. Collateral circulation in symptomatic intracranial atherosclerosis. J Cereb Blood Flow Metab. 2011;31:1293–1301.

49. Mangiardi M, Bonura A, Iaccarino G, Alessiani M, Bravi MC, Crupi D, Pezzella FR, Fabiano S, Pampana E, Stilo F, et al. The Pathophysiology of Collateral Circulation in Acute Ischemic Stroke. Diagnostics (Basel). 2023;13:2425.

50. Guglielmi V, LeCouffe NE, Zinkstok SM, Compagne KCJ, Eker R, Treurniet KM, Tolhuisen Manon L, van der Worp HB, Jansen IGH, van Oostenbrugge RJ, et al. Collateral Circulation and Outcome in Atherosclerotic Versus Cardioembolic Cerebral Large Vessel Occlusion. Stroke. 2019;50:3360–3368.

51. Izumi C, Miyake M, Amano M, Matsutani H, Hashiwada S, Kuwano K, Kuroda M, Nishimura S, Yoshikawa Y, Takahashi Y, et al. Risk Factors of Aortic Plaque Progression Evaluated by Long-Term Follow-Up Data With Transesophageal Echocardiography. The American Journal of Cardiology. 2017;119:1872–1876.

52. Camaj A, Fuster V, Giustino G, Bienstock SW, Sternheim D, Mehran R, Dangas GD, Kini A, Sharma SK, Halperin J, et al. Left Ventricular Thrombus Following Acute Myocardial Infarction: JACC State-of-the-Art Review. J Am Coll Cardiol. 2022;79:1010–1022.

53. Cisneros S, Duarte R, Fernandez-Perez GC, Castellon D, Calatayud J, Lecumberri I, Larrazabal E, Ruiz BI. Left ventricular apical diseases. Insights Imaging. 2011;2:471–482.

54. Gottdiener JS, Massie B, Ammons SB, Egher C, Petillo F, Krol WF, Horney RN, Collins JF. Prevalence of left ventricular thrombus in dilated cardiomyopathy: The WATCH trial. Journal of the American College of Cardiology. 2003;41:202–202.

55. Eberli FR, O’Sullivan CJ. Left ventricular thrombus formation after acute myocardial infarction: vigilance still required in the modern era. Swiss Medical Weekly. 2015;145:w14158–w14158.

56. Levine GN, McEvoy JW, Fang JC, Ibeh C, McCarthy CP, Misra A, Shah ZI, Shenoy C, Spinler SA, Vallurupalli S, et al. Management of Patients at Risk for and With Left Ventricular Thrombus: A Scientific Statement From the American Heart Association. Circulation. 2022;146:e205–e223.

## References

18. Ntaios G, Weng SF, Perlepe K, Akyea R, Condon L, Lambrou D, Sirimarco G, Strambo D, Eskandari A, Karagkiozi E, et al. Data-driven machine-learning analysis of potential embolic sources in embolic stroke of undetermined source. European Journal of Neurology. 2021;28:192–201.

19. Kamel H, Navi BB, Parikh NS, Merkler AE, Okin PM, Devereux RB, Weinsaft JW, Kim J, Cheung JW, Kim LK, et al. Machine Learning Prediction of Stroke Mechanism in Embolic Strokes of Undetermined Source. Stroke. 2020;51:e203–e210.

20. Saito T, Rehmsmeier M. The precision-recall plot is more informative than the ROC plot when evaluating binary classifiers on imbalanced datasets. PLoS One. 2015;10:e0118432.

21. Owusu-Adjei M, Hayfron-Acquah JB, Frimpong T, Abdul-Salaam G. Imbalanced class distribution and performance evaluation metrics: A systematic review of prediction accuracy for determining model performance in healthcare systems. PLOS Digital Health. 2023;2:e0000290.

22. Speiser JL, Kerr WT, Ziegler A. Common Critiques and Recommendations for Studies in Neurology Using Machine Learning Methods. Neurology. 2024;103:e209861.

23. Sanfilippo AJ, Abascal VM, Sheehan M, Oertel LB, Harrigan P, Hughes RA, Weyman AE. Atrial enlargement as a consequence of atrial fibrillation. A prospective echocardiographic study. Circulation. 1990;82:792–797.

24. Zacà V, Galderisi M, Mondillo S, Focardi M, Ballo P, Guerrini F. Left atrial enlargement as a predictor of recurrences in lone paroxysmal atrial fibrillation. Can J Cardiol. 2007;23:869–872.

25. Abhayaratna WP, Seward JB, Appleton CP, Douglas PS, Oh JK, Tajik AJ, Tsang TSM. Left Atrial Size: Physiologic Determinants and Clinical Applications. Journal of the American College of Cardiology. 2006;47:2357–2363.

26. Jordan K, Yaghi S, Poppas A, Chang AD, Mac Grory B, Cutting S, Burton T, Jayaraman M, Tsivgoulis G, Sabeh MK, et al. Left Atrial Volume Index Is Associated With Cardioembolic Stroke and Atrial Fibrillation Detection After Embolic Stroke of Undetermined Source. Stroke. 2019;50:1997–2001.

27. Thomas L, Levett K, Boyd A, Leung DYC, Schiller NB, Ross DL. Compensatory changes in atrial volumes with normal aging: is atrial enlargement inevitable? Journal of the American College of Cardiology. 2002;40:1630–1635.

28. Aurigemma GP, Gottdiener JS, Arnold AM, Chinali M, Hill JC, Kitzman D. Left atrial volume and geometry in healthy aging: the Cardiovascular Health Study. Circ Cardiovasc Imaging. 2009;2:282–289.

29. Prastaro M, Paolillo S, Savarese G, Dellegrottaglie S, Scala O, Ruggiero D, Gargiulo P, Marciano C, Parente A, Cecere M, et al. N-terminal pro-b-type natriuretic peptide and left atrial function in patients with congestive heart failure and severely reduced ejection fraction†. European Journal of Echocardiography. 2011;12:506–513.

30. Tsuchida K, Tanabe K. Influence of paroxysmal atrial fibrillation attack on brain natriuretic peptide secretion. J Cardiol. 2004;44:1–11.

31. Seegers J, Zabel M, Grüter T, Ammermann A, Weber-Krüger M, Edelmann F, Gelbrich G, Binder L, Herrmann-Lingen C, Gröschel K, et al. Natriuretic peptides for the detection of paroxysmal atrial fibrillation. Open Heart [Internet]. 2015 [cited 2025 Feb 13];2. Available from: https://openheart.bmj.com/content/2/1/e000182

32. di Biase L, Bonura A, Pecoraro PM, Carbone SP, Di Lazzaro V. Unlocking the Potential of Stroke Blood Biomarkers: Early Diagnosis, Ischemic vs. Haemorrhagic Differentiation and Haemorrhagic Transformation Risk: A Comprehensive Review. Int J Mol Sci. 2023;24:11545.

33. Bar SL, Swiggum E, Straatman L, Ignaszewski A. Nonheart failure-associated elevation of amino terminal pro-brain natriuretic peptide in the setting of sepsis. Can J Cardiol. 2006;22:263–266.

34. Inoue S-I, Murakami Y, Sano K, Katoh H, Shimada T. Atrium as a source of brain natriuretic polypeptide in patients with atrial fibrillation. Journal of Cardiac Failure. 2000;6:92–96.

35. Annoura M, Ogawa M, Kumagai K, Zhang B, Saku K, Arakawa K. Cholesterol paradox in patients with paroxysmal atrial fibrillation. Cardiology. 1999;92:21–27.

36. Lee H, Lee S, Choi E, Han K, Oh S. Low Lipid Levels and High Variability are Associated With the Risk of New-Onset Atrial Fibrillation. Journal of the American Heart Association. 2019;8:e012771.

37. Hanna IR, Heeke B, Bush H, Brosius L, King-Hageman D, Dudley SC, Beshai JF, Langberg JJ. Lipid-lowering drug use is associated with reduced prevalence of atrial fibrillation in patients with left ventricular systolic dysfunction. Heart Rhythm. 2006;3:881–886.

38. Goonasekara CL, Balse E, Hatem S, Steele DF, Fedida D. Cholesterol and cardiac arrhythmias. Expert Rev Cardiovasc Ther. 2010;8:965–979.

39. Guo Y, Lip GYH, Apostolakis S. Inflammation in atrial fibrillation. J Am Coll Cardiol. 2012;60:2263–2270.

40. Krishnan A, Sharma H, Yuan D, Trollope AF, Chilton L. The Role of Epicardial Adipose Tissue in the Development of Atrial Fibrillation, Coronary Artery Disease and Chronic Heart Failure in the Context of Obesity and Type 2 Diabetes Mellitus: A Narrative Review. J Cardiovasc Dev Dis. 2022;9:217.

41. Kamel H, Healey JS. Cardioembolic Stroke. Circ Res. 2017;120:514–526.

42. Lin HJ, Wolf PA, Kelly-Hayes M, Beiser AS, Kase CS, Benjamin EJ, D’Agostino RB. Stroke severity in atrial fibrillation. The Framingham Study. Stroke. 1996;27:1760–1764.

43. Amarenco P, Cohen A, Tzourio C, Bertrand B, Hommel M, Besson G, Chauvel C, Touboul PJ, Bousser MG. Atherosclerotic disease of the aortic arch and the risk of ischemic stroke. N Engl J Med. 1994;331:1474–1479.

44. Jones EF, Kalman JM, Calafiore P, Tonkin AM, Donnan GA. Proximal aortic atheroma. An independent risk factor for cerebral ischemia. Stroke. 1995;26:218–224.

45. Harloff A, Simon J, Brendecke S, Assefa D, Helbing T, Frydrychowicz A, Weber J, Olschewski M, Strecker C, Hennig J, et al. Complex plaques in the proximal descending aorta: an underestimated embolic source of stroke. Stroke. 2010;41:1145–1150.

46. Liebeskind DS, Cotsonis GA, Saver JL, Lynn MJ, Cloft HJ, Chimowitz MI, Warfarin–Aspirin Symptomatic Intracranial Disease (WASID) Investigators. Collateral circulation in symptomatic intracranial atherosclerosis. J Cereb Blood Flow Metab. 2011;31:1293–1301.

47. Mangiardi M, Bonura A, Iaccarino G, Alessiani M, Bravi MC, Crupi D, Pezzella FR, Fabiano S, Pampana E, Stilo F, et al. The Pathophysiology of Collateral Circulation in Acute Ischemic Stroke. Diagnostics (Basel). 2023;13:2425.

48. Guglielmi V, LeCouffe NE, Zinkstok SM, Compagne KCJ, Eker R, Treurniet KM, Tolhuisen Manon L, van der Worp HB, Jansen IGH, van Oostenbrugge RJ, et al. Collateral Circulation and Outcome in Atherosclerotic Versus Cardioembolic Cerebral Large Vessel Occlusion. Stroke. 2019;50:3360–3368.

49. Izumi C, Miyake M, Amano M, Matsutani H, Hashiwada S, Kuwano K, Kuroda M, Nishimura S, Yoshikawa Y, Takahashi Y, et al. Risk Factors of Aortic Plaque Progression Evaluated by Long-Term Follow-Up Data With Transesophageal Echocardiography. The American Journal of Cardiology. 2017;119:1872–1876.

50. Camaj A, Fuster V, Giustino G, Bienstock SW, Sternheim D, Mehran R, Dangas GD, Kini A, Sharma SK, Halperin J, et al. Left Ventricular Thrombus Following Acute Myocardial Infarction: JACC State-of-the-Art Review. J Am Coll Cardiol. 2022;79:1010–1022.

51. Cisneros S, Duarte R, Fernandez-Perez GC, Castellon D, Calatayud J, Lecumberri I, Larrazabal E, Ruiz BI. Left ventricular apical diseases. Insights Imaging. 2011;2:471–482.

52. Gottdiener JS, Massie B, Ammons SB, Egher C, Petillo F, Krol WF, Horney RN, Collins JF. Prevalence of left ventricular thrombus in dilated cardiomyopathy: The WATCH trial. Journal of the American College of Cardiology. 2003;41:202–202.

